# Early recovery during stroke rehabilitation predicts the functional outcome at discharge: a retrospective clinical study

**DOI:** 10.1101/2025.02.03.25321578

**Authors:** Marco Anziano, Riccardo Cusinato, Athina Tzovara, Lucas Spierer, Joelle N. Chabwine

## Abstract

**Background:** Models predicting functional recovery upon stroke rehabilitation (SR) are not implemented in clinical practice due to their complexity and limited power in patients with moderate/severe deficits.

**Objectives:** To obtain preliminary evidence supporting the predictive value of a single measure of early recovery on SR discharge functional status.

**Methods:** We tested simple predictive models based on the functional independence measure (FIM) changes during the first three weeks of SR (3wFIMgain) in 81 patients with moderate/severe impairment.

**Results:** Patients with poor or good 3wFIMgain featured similar admission impairment, rate of complications, presence of aphasia or dominant-sided hemisyndrome, while poor early recovery was associated with lesions to the left sensory-motor cortex, temporo-parietal junction, and superior longitudinal fasciculus.

The 3wFIMgain effectively predicted the discharge FIM gain in patients with ≥ 6 weeks of SR stay. Combined with the admission FIM, the 3wFIMgain improved a predictive model for the discharge FIM.

**Conclusions:** The 3wFIMgain bears strong discharge prognostic value and synthetizes complex information about the early recovery of stroke patients that cannot be described by individual clinical predictors. Measures of early recovery might improve the power of models and simplify them, thus favoring application in clinical practice. Future validation in confirmatory studies is required.

## 1. Introduction

Predicting the functional outcome of patients following inpatient stroke rehabilitation (SR) is crucial for appropriate therapies planning, to identify the most adapted discharge destination, as well as to clearly inform patients and their relatives about post-stroke recovery prognosis^1,2^. Although current prognostic models provide good estimates of group functional outcome^1,3,4^, their translation into clinical practice fails plausibly because i) they are not accurate enough to predict individual outcomes, especially in severely impaired patients on admission to SR, and ii) they remain mathematically complex and require a large amount of patient-related information to be applicable^5–7^. Here, we tested the hypothesis that simpler prognostic models based on measures of patients’ recovery during early stages of SR (hereafter referred to as “early recovery”) could solve the above-mentioned issues. Indeed, because early recovery results from interactions between many clinical factors, we posit that synthetizing such information in a single predictor would improve the prediction of both the functional independence and total gain at discharge, including for patients with severe initial impairment. Predicting the total gain is important because it expresses how much the patient had improved regardless of his/her initial disability and is directly related to long-term mortality^8^. In addition, simplifying prognostic models by using this readily available information would promote their use by health professionals as a practical tool to predict individual recovery potential and refine rehabilitation management plans.

The few existing studies focusing on the predictive power of early recovery suggest that adding such predictors produces models that outperform those based solely on information available on admission to SR, even though this gain in performance remains at the expense of additional data to be collected up to four to six weeks after admission^9,10^.

As a proof of concept, we conducted a monocentric retrospective study of stroke patients with moderate to severe functional impairment to obtain preliminary data elucidating whether simple models based on the functional independence measure (FIM) gain during the first three weeks of rehabilitation (3wFIMgain) would detect patients with large functional recovery and predict patients’ individual values of both discharge FIM (DFIM) and DFIM gain. This approach allowed us to test the validity of the 3wFIMgain to support patient-specific SR planning by identifying patients with good recovery potential, thereby refining prognostic estimates.

To further examine the determinants of early recovery we searched for clinical factors associated with patients showing poor early recovery (PER). As a supplementary analysis, we performed lesion-symptom mapping to identify brain structures whose damage was associated with PER. We expected the patients’ early recovery to be independent from their impairment on admission, whereas PER to be associated with specific clinical characteristics (higher rate of complications and level of comorbidities, presence of aphasia/dominant-lateralized hemisyndrome, etc.) and lesions to brain structures responsible for major neuropsychological or sensory-motor syndromes.

## 2. Materials and methods

The study was approved by the Research Ethics Committee of the Canton de Vaud, Switzerland (project-ID 2018-01895) and was conducted in compliance with the Helsinki Declaration^11^ and other international ethical rules. All reachable participants (or their legal proxies) gave their informed consent to use their data.

### 2.1 Participants

We retrospectively collected information on stroke patients admitted in the Neurorehabilitation Division of the Fribourg Hospital, site of Meyriez (Switzerland) between 2017 and 2019.

Inclusion criteria were: i) first ischemic or hemorrhagic symptomatic stroke; ii) total FIM score on admission (AFIM) <100; iii) available brain CT or MRI images from routine stroke assessment. Exclusion criteria were: i) medical history of major central nervous system or psychiatric pathologies ii) existence of other symptomatic neurological or psychiatric disorder potentially interfering with the physical or neuropsychological recovery; iii) Premorbid modified rankin scale (MRS, ^12^) ≥ 2.

A total of 89 patients were selected for the study. The analyses were limited to patients having a minimum length of stay (LOS) of three weeks (N=84), of which three were excluded during the clustering procedure (see paragraph 2.3.1), leaving a final sample of 81 patients finally included. A subgroup of 49 patients having a LOS ≥ 6 weeks was selected for the analyses of predictive validity (see paragraph 2.3). Demographic and clinical-related data are reported in table 1.

**Table 1.** Demographic and clinical-related data of the whole cohort and separated according to the amount of FIM gain obtained in the first three weeks of rehabilitation.

|  | Whole cohort (N=81) |  | Poor early recovery (N=46) |  | Good early recovery (N=35) |  | good vs poor |
| --- | --- | --- | --- | --- | --- | --- | --- |
|  | Mean (SD) or Number | Range | Mean (SD) or Number | Range | Mean (SD) or Number | Range | p value (BF<sub>01</sub>) |
| Age (years) | 73.2 (12.0) | 42-97 | 71.7 (13.2) | 42-97 | 75.2 (10.0) | 46-89 | 0.19 |
| Gender (M/F) | 43/38 | - | 27/19 | - | 16/19 | - | - |
| Ischemic/ Hemorrhagic stroke | 63/18 | - | 36/10 | - | 27/8 | - | - |
| Admission FIM | 60.6 (25.3) | 18-99 | 57.0 (28.2) | 18-99 | 65.3 (20.4) | 26-96 | 0.17 (2.2) |
| Discharge FIM | 93.5 (27.6) | 22-125 | 81.5 (30.2) | 22-121 | 109.3 (11.6) | 69-125 | **<0.01\*** |
| Discharge FIM gain | 32.9 (18.4) | -1-82 | 24.5 (15.8) | -1-67 | 44.0 (15.7) | 18-82 | **<0.01\*** |
| 3 weeks FIM gain | 17.2 (11.2) | -3-49 | 9.6 (6.0) | -3-22 | 27.3 (8.0) | 10-49 | **<0.01** |
| Admission FIM cognitive | 18.7 (7.8) | 5-35 | 17.3 (8.6) | 5-33 | 20.6 (6.3) | 9-35 | 0.12 |
| Onset NIHSS | 11.5 (7.2) | 0-28 | 12.2 (6.3) | 3-25 | 10.7 (8.1) | 0-28 | 0.45 (4.0) |
| Onset-admission delay (days) | 21.1 (16.8) | 1-119 | 24.7 (19.6) | 1-119 | 18.9 (11.5) | 2-51 | 0.17 |
| Length of stay (days) | 60.7 (34.4) | 12-158 | 67.8 (36.2) | 20-158 | 51.3 (29.8) | 12-124 | - |
| Admission CIRS<sup>†</sup> | 13.8 (7.1) | 2-29 | 14.7 (7.1) | 3-28 | 12.6 (7.0) | 2-29 | 0.19 |
| N. of complications (0/1/ 2/ ≥ 3) | 40/19/9/13 | - | 22/9/6/9 | - | 18/10/3/4 | - | 0.59<sup>x</sup> |
| N. of complications during the first 3 weeks of rehabilitation | 17 | - | 11 | - | 6 | - | - |
| Aphasia/dominant hemisindrome (no/yes) | 35/46 | - | 17/29 | - | 18/17 | - | 0.28<sup>x</sup> |

BF<sub>01</sub>: Bayes factor in favor of the null hypothesis; CIRS: Cumulative Illness Rating Scale; FIM: Functional Independence Measure; NIHSS: National Institutes of Health Stroke Scale; SD: Standard Deviation. † non-neurological comorbidities. \* Independent samples test conducted on the subgroup of 49 patients with length of stay ≥ 6 weeks. <sup>x</sup> Chi-square test. Significant P values are in bold. NIHSS was available for 61/81 patients.

### 2.2 The functional independence measures

The FIM, adopted as the main functional measure in this study, is a measure of independence in the basic activities of daily life, displaying overall good psychometric characteristics^13,14^, and widely used in rehabilitation settings. It is an ordinal scale featuring 18 items for motor and cognitive domains, each valued from 1 to 7 with increasing values expressing better levels of independence in a single activity (1=total dependence; 7 = total independence, range=18-126). In the present study, we focused on the total FIM score. Specifically, we used the AFIM and the FIM gain during the first three weeks of SR (3wFIMgain) as independent variables; and the discharge FIM (DFIM) and the DFIM gain as dependent variables. The 3wFIMgain was calculated as the FIM score at three weeks of SR minus the AFIM. The DFIM gain was calculated as the DFIM minus the AFIM.

### 2.3 Statistical analyses

We performed statistical inferential tests (correlations, independent group tests, linear regressions, Chi-square. We additionally conducted Bayes factor (BF) analyses with a prior of 1 where we expected no difference between the groups. When relevant violations of the statistic assumption were estimated, we adopted corresponding nonparametric approaches for the analyses (Mann-Whitney U for independent group tests, spearman correlations). Correction for family-wise error rate (FWER) within each research question was conducted with Bonferroni-Holmes method. These analyses were performed with the software Jamovi version 1.2 ^15^.

We focused on patients’ 3wFIMgain as a predictor because a three week period offered a sufficient number of time points to define characteristic profiles of recovery (see paragraph 2.3.1), and allowed to include the vast majority of patients in the analyses (84 out of 89). A three-weeks period also constituted a valid delay for prognostic evaluation in patients with moderate to severe deficits for whom duration of stay in SR commonly exceeds this length in many countries worldwide^16–19^.

However, because the 3wFIMgain expresses a variable portion of the DFIM gain depending on the LOS, both FIM gains were intrinsically correlated and even coincide in case of LOS = 3 weeks. To compensate for this aspect all prediction or correlation analyses for the outcomes based on DFIM and DFIM gain (see the paragraphs 2.3.3 and 2.3.4) were limited to a subgroup of 49 patients with a LOS ≥ 6 weeks, the majority of which had a LOS superior to 8 weeks (1st quartile LOS = 57 days). The association of the AFIM and the National Institutes of Health Stroke Scale (NIHSS^20^; a widely adopted 11 items scale of stroke-induced neurological deficit ranging from 0 to 42 points, higher scores corresponding to more severe deficit) with both DFIM and DFIM gain was also analyzed in this subgroup of 49 patients to allow for adequate comparisons of the results (paragraphs 2.3.3 and 2.3.4).

#### 2.3.1. Definition of the early recovery profiles

To define the patients’ early recovery profiles, we grouped patients with similar weekly FIM evolution trajectories using unsupervised learning (clustering). To this aim, we focused on the patients’ FIM values on admission and during the subsequent three weeks, extracting the resulting three values of weekly FIM gain, (Week1-Admission, Week2-Week1 and Week3-Week-2).

Prior to the clustering procedure, we ran an outlier detection algorithm based on the Mahalanobis distance^21^ with probability threshold set at 1%. This standard outliers’ removal procedure allows avoiding distorted clusters by identifying samples (patients) that present an abnormal distance from all the other samples in the feature space (weekly FIM gain). Three patients were excluded at this step from the initial sample of N=84 (see paragraph 2.1).

We then employed K-means clustering, a widely used algorithm that iteratively searches for the center of each cluster based on Euclidean distances^22,23^ (default SciKit-learn implementation^24^). We chose the number of clusters (k=3) based on standard metrics in the field (within-cluster sum of squares, silhouette score, Calinski-Harabasz Index, Davies-Bouldin index) and because this value was clinically meaningful, as the resulting clusters represented three clearly distinct profiles of evolution during the first three weeks of SR (figure 1).

**Figure 1.**
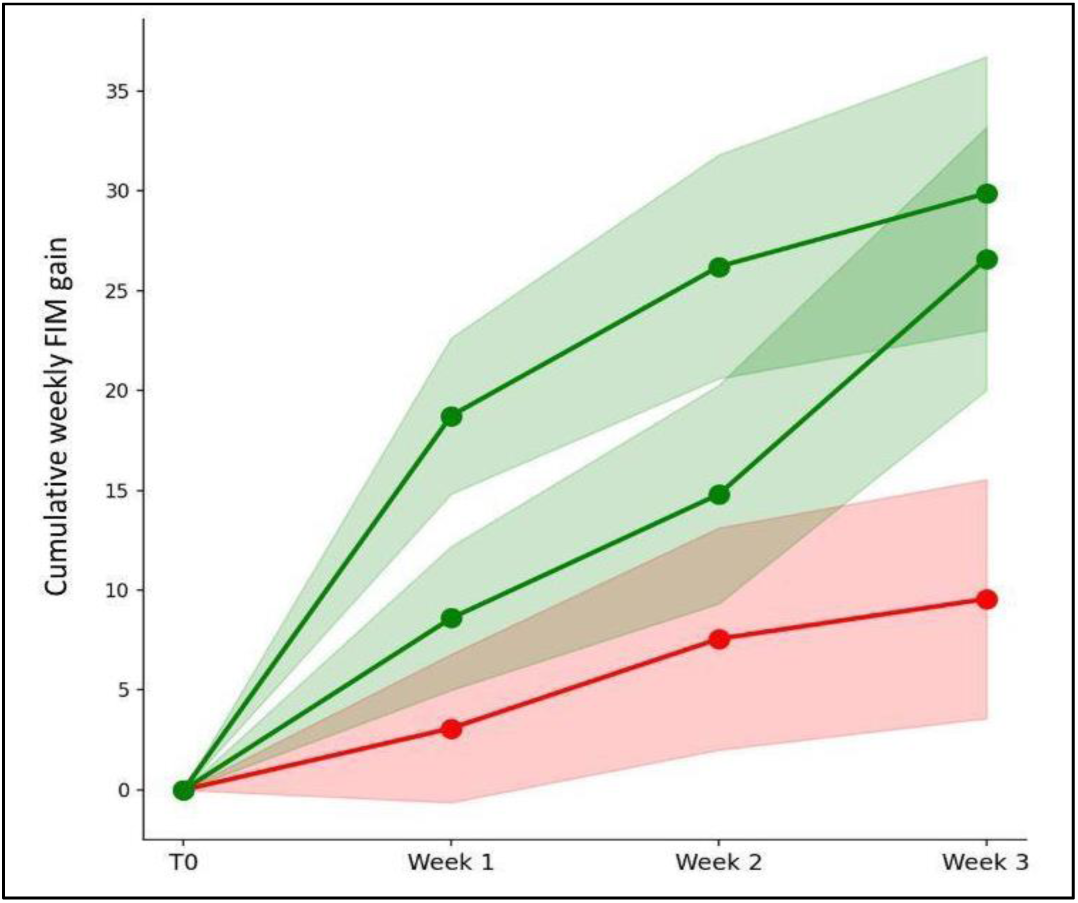
Early recovery profiles based on weekly FIM scores (clusters): X axis, weeks from admission to inpatient rehabilitation; Y axis, cumulative values of the weekly FIM gain during the first three weeks of rehabilitation (e.g., cumulative FIM gain at week 2 = FIM gain at week1 (Week1-T0) + FIM gain at Week2 (Week2-Week1)) Lines represent the average values of cumulative weekly FIM gain of each cluster. Standard deviation is shown as shaded areas. Red cluster (N=46); Middle green cluster(N=20); Top green cluster (N=15). Patients from the two green clusters have been unified to constitute the group of patients with good early recovery, while the red cluster corresponds to the group of patients with poor early recovery.

Because the two clusters with good recovery presented on average similar values of 3wFIMgain, despite displaying a different profile of evolution (green curves in figure 1), we unified their data to increase our statistical power. The resulting group represented the final category of patients with good early recovery (GER) and we conducted multiple analyses confronting this group with the cluster of poor early recovery (PER, red curve in figure 1), featuring the lowest average 3wFIMgain.

Similarly, to perform correlations, linear regressions, and lesion-symptom mapping (see paragraph 2.3.4 and 2.3.5) analyses, the Euclidean distance of each individual patient’s profile from the center of the PER cluster was used. This continuous variable expresses how much the single patient’s early evolution trajectory differs from the average profile of the PER.

#### 2.3.2 Clinical factors characterizing the early recovery profiles

We explored whether a set of clinical factors could influence early recovery and therefore be significantly different between the two groups of GER and PER. We analyzed: i) NIHSS at stroke onset; ii) AFIM; iii) Age; iv) non-neurological comorbidities on admission measured by the cumulative illness rating scale (CIRS25); v) presence of aphasia and/or dominant-sided sensory-motor hemisyndrome (yes/no); vi) cognitive FIM subcomponent on admission (AFIMc) vii) number of complications (three categories: 1, 2, ≥ 3); viii) stroke onset-SR admission delay (in days).

The CIRS is a validated cumulative score measuring the patient’s overall burden of disease. It features a 0 to 4 score (0= no alteration; 4=Acute organ insufficiency requiring emergency therapy) to define the degree of alteration of each of 14 physiological systems^25^. To specifically evaluate the impact of comorbidities not directly related to the stroke, we excluded the item relative to the nervous system, producing a derivative score ranging from 0 to 52. We considered as complication, any major clinical condition occurring during a period spanning from the stroke onset to the first three weeks of SR (including the acute post-stroke hospitalization) and causing a considerable degradation of the patient’s general condition (major infections, heart decompensation, intracranial hypertension with craniotomy, need for mechanical ventilation, etc.) or interfering with rehabilitative interventions (fractures, musculoskeletal affections, severe pain-related problems, etc.). Aphasia and dominant hemisyndrome as well as the AFIMc were chosen based on the results of our lesion-symptom mapping analysis revealing a possible relationship between bad evolution and lesions located in left-sided brain regions, typically associated with these syndromes or impacting the cognitive domain (see paragraph 2.3.5).

For continuous variables we performed Welch’s t-tests or non-parametric Mann-Whitney U tests. For the two categorical variables (complications, Aphasia/ dominant hemisyndrome) we performed Chi-square tests. These analyses were conducted on the full cohort of 81 patients.

#### 2.3.3 Association of early recovery profiles with outcomes

To assess the applicability of FIM profiles as predictors of functional outcome (analyses conducted on the subgroup with LOS ≥ 6 weeks; N=49; see paragraph 2.3), we first tested whether the GER and PER groups differed in terms of DFIM and DFIM gain through independent group tests. The DFIM and DFIM gain express respectively the final functional status and the overall amount of functional recovery obtained during the SR. We further investigated correlations of DFIM and DFIM gain with the Euclidean distance of the individual patient’s profile from the center of the PER cluster. A positive correlation would mean that the more the patient’s profile differs from that of the PER group, the better would be his/her outcome.

For comparison with the above-mentioned analyses, we tested in the same subgroup correlations of commonly adopted predictors such as AFIM and NIHSS at stroke onset, with DFIM and DFIM gain. The results of these analyses are presented in the supplementary material and mentioned in the discussion.

#### 2.4.4 Predictive value of early FIM gain

We found a strong positive correlation between the Euclidean distance from the center of the PER cluster and the 3wFIMgain (Pearson’s r= 0.86, p(81)<0.01), which confirmed that the 3wFIMgain constitutes a simple and effective way to synthesize the separation of patients in the GER and PER outcome groups. Given the greater practicality of using the 3wFIMgain in clinical settings compared to cluster analyses, we focused on this parameter for the prognostic application in SR.

We tested the 3wFIMgain predictive power for DFIM gain using two statistical methods: i) we calculated the receiver operating characteristic curve (ROC) for the categorization of patients using the software Jamovi version 1.2^15^. Patients were separated into two outcome groups of poor/good total recovery based on the cutoff value of DFIM gain>49 points. This value, although arbitrary, represents a clinically remarkable amount of recovery corresponding to more than two times the “minimal clinical important difference” (MCID) for the FIM score^26^. We then computed the area under the curve (AUC) and the cutoff for the 3wFIMgain score featuring optimal balance between sensitivity and specificity. ii) we modelled a simple linear regression between these two variables to test the validity of 3wFIMgain in predicting the DFIM gain individual scores.

Moreover, given the importance of AFIM as a predictor of functional outcome and our confirmation of its good correlations of this factor with DFIM (see supplementary material), we combined AFIM and 3wFIMgain in a linear model to investigate whether the addition of 3wFIMgain as dependent variable would improve the overall predictive power of the model for the DFIM. These analyses were conducted on the subgroup with LOS > 6 weeks (N=49).

#### 2.3.5 Voxel-wise lesion-symptom mapping

To explore whether specific stroke lesion locations were associated with poor early recovery we conducted a Voxel-wise lesion-symptom mapping (VLSM) analysis on the Euclidean distance of the individual patient’s profile from the center of the PER cluster.

We retrospectively extracted structural CT/MRI (23/58) brain images acquired as part of the routine clinical investigations from the database of the Fribourg Hospital, Switzerland. Lesion demarcation and normalization were performed with a manual approach^27^. Masks were drawn directly on the SPM12-provided MNI152 Nonlinear (2009) brain template^28^ using the software MRIcron^29^, procedure detailed in Chouiter et al. 2016^30^.

We conducted the VLSM analysis using the software Niistat (procedure described in Niistat, URL: http://www.nitrc.org/plugins/mwiki/index.php/niistat^31^). To obtain an optimal balance between brain coverage and sufficient statistical power we analyzed only voxels that were damaged in at least six patients. We adopted a one-tailed 0.05 alpha threshold with a permutation-based correction for FWER with 5000 permutations.

## 3. Results

### 3.1 Clinical factors characterizing the early recovery profiles

Clinical-related data and results for the comparison tests between the GER (N=35) and PER (N=46) groups are reported in table 1. The two groups showed no significant difference for AFIMc (Mann-Whitney U test; p=0.12, mean difference=3.3, good>poor), stroke onset-admission delay (Mann-Whitney U test; p=0.17, mean difference=5.8, poor>good), CIRS (Welch’s t; p(74.0)=0.19, Cohen’s d=0.30, poor>good), and age (Welch’s t; p(79.0)=0.19, Cohen’s d=-0.29, poor>good). We also found no difference for the AFIM (Mann-Whitney U test; p=0.17, mean difference=8.3, good>poor) and NIHSS (Welch’s t; p(52.9)=0.45, Cohen’s d=0.20, poor>good), with BF_01_ of 2.2 and 4.0 respectively, meaning negligible positive evidence in favor of the null hypothesis. The two groups showed no significant difference also for the number of complications (X2, p(3)=0.59) or the presence of Aphasia and/or dominant hemisyndrome (X2 with continuity correction, p(1)=0.28) with slightly smaller occurrence of Aphasia and/or dominant hemisyndrome in the GER group (Odds ratio= 0.55, 95%CI, 0.23-1.35).

### 3.2 Predictive value of early FIM gain

Prediction analyses were performed on the subgroup of 49 patients with a LOS ≥ 6 weeks. The cutoff of 3wFIMgain>19 points was the best to classify the patients based on their DFIM gain and the calculated AUC of the ROC was 0.858. A 3wFIMgain score >19 points correctly predicted a good total recovery (DFIM gain>49) in 83.3% of the cases (sensitivity=10/12), while inferior scores of 3wFIMgain correctly predicted a poor total recovery in 81.1% of the cases (specificity= 30/37).

The 3wFIMgain presented a significant positive linear relationship with the DFIM gain, explaining 44% of its variance (p(1,47)<0.01, adjusted R^2^=0.436) with a root mean squared error (RMSE) of 14.4 points of DFIM gain (figures 2a and 2b). Specifically, an increase of 1 point in 3wFIMgain predicted an increase of 1.12 points of DFIM gain (p<0.01, 95%CI: 0.45-0.89).

**Figure 2.**
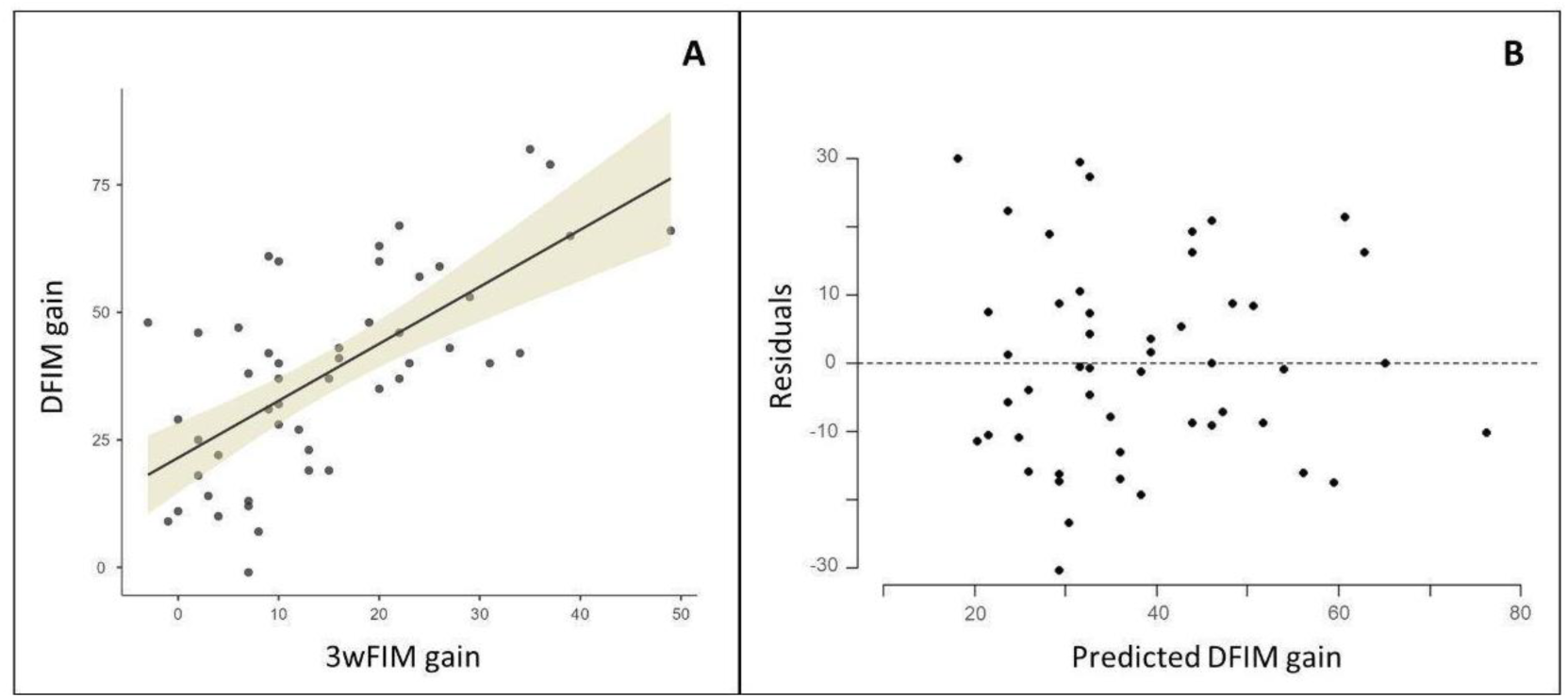
Predictive value of early FIM gain for DFIM gain. A: Linear model of 3wFIMgain explaining variations in DFIM gain in the subgroup of patients with a minimal stay of 6 weeks (N=49); Shaded area represents 95% confidence intervals for the predicted mean value of DFIM gain. B: Residual plot of the model.

The AFIM showed a significant positive linear relation with the DFIM, explaining 57% of its variance (p(1,47)<0.01, adjusted R^2^=0.566) with a RMSE of 19.4 points of DFIM (figures 3a and 3b). An increase of 1 point in AFIM predicted an increase of 0.97 points in DFIM (p<0.01, 95%CI: 0.57-0.95). The addition of the 3wFIMgain as a second independent variable significantly improved the model (p(1,46)<0.01, ΔR^2^=0.207) that was now able to explain 77% of the variance in DFIM (p(2,46)<0.01, adjusted R^2^=0.772) with a RMSE of 13.9 points of DFIM (figure 3c). Both variables were significant predictors with a greater standardized coefficient for AFIM (AFIM stand. estimate=0.65, p<0.01; 3wFIMgain stand. estimate=0.47, p<0.01). The equations derived from the linear models for the prediction of DFIM and DFIM gain are shown in table 2.

**Figure 3.**
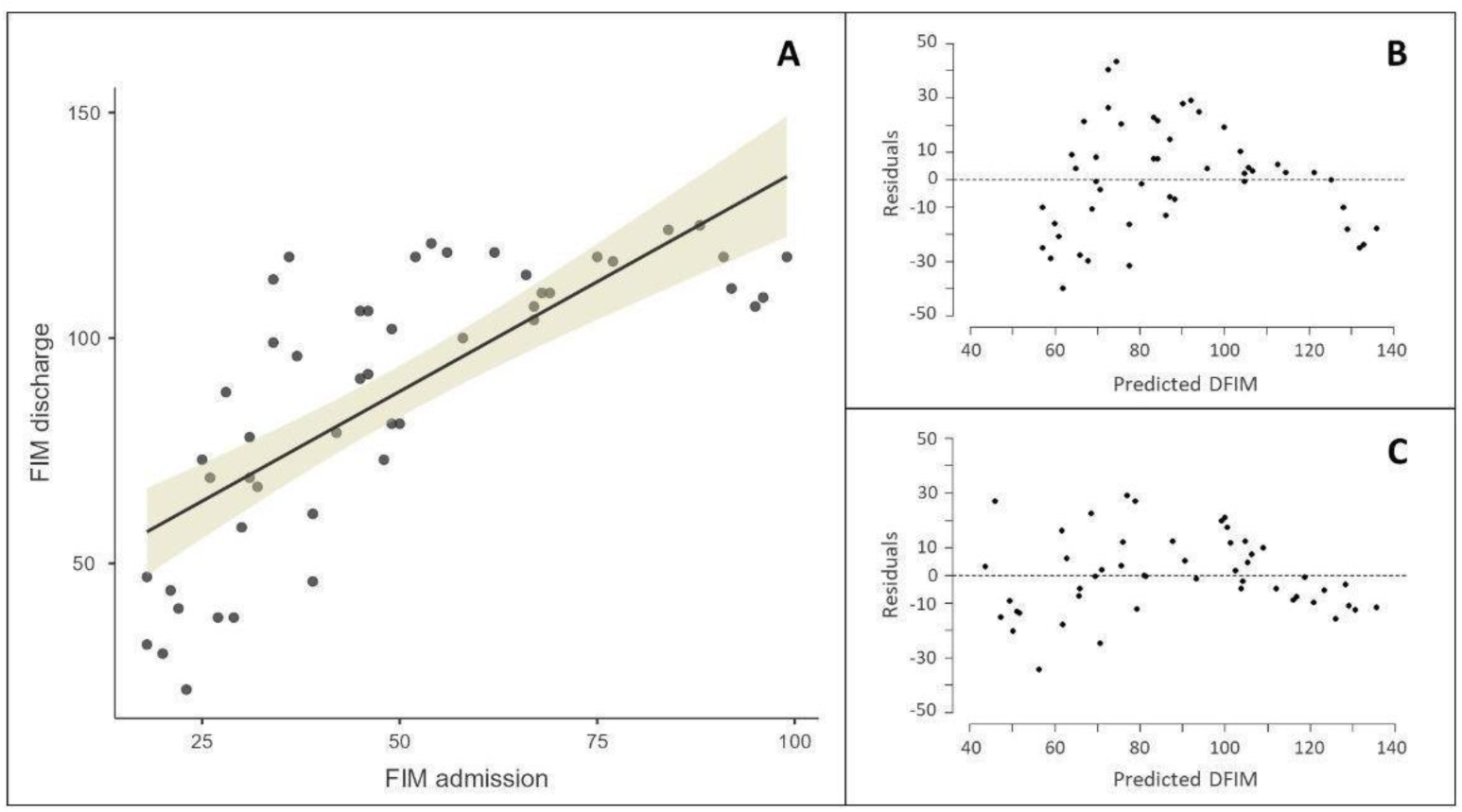
Early FIM gain improves the predictive value of AFIM for DFIM. A: Simple linear model of AFIM explaining variations in DFIM in the subgroup of patients with a minimal stay of 6 weeks (N=49); Shaded area represents 95% confidence intervals for the predicted mean value of DFIM. B: Residual plot of the simple linear model presented in A. C: Residual plot of the model with the two predictors AFIM and 3wFIMgain explaining variations in DFIM in the subgroup of patients with a minimal stay of 6 weeks.

**Table 2.**
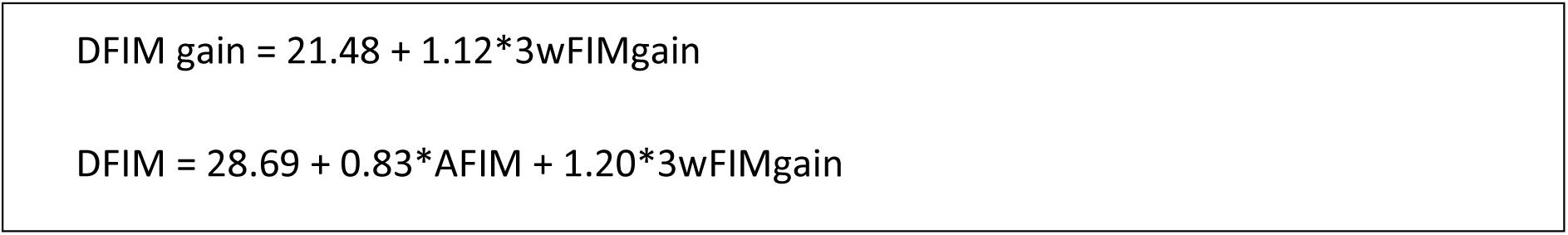
Equations for the prediction of functional outcomes on discharge after inpatient neurorehabilitation.

### 3.3 Voxel-wise lesion-symptom mapping

The map of brain lesion coverage is shown in figure 4a. Although no association with poor recovery was significant after correcting for the FWER, three clusters localized in the left hemisphere (figure 4b), displayed near-threshold Z-scores and were situated in the: i) white matter deeply to the temporo-parietal junction, mainly intersecting the superior longitudinal fasciculus (MNIxyz coordinates –33, –30, 29); ii) supramarginal gyrus cortex and subcortex (MNIxyz coordinates –59, –26, 23); iii) precentral gyrus (MNIxyz coordinates –40, –7, 43).

**Figure 4.**
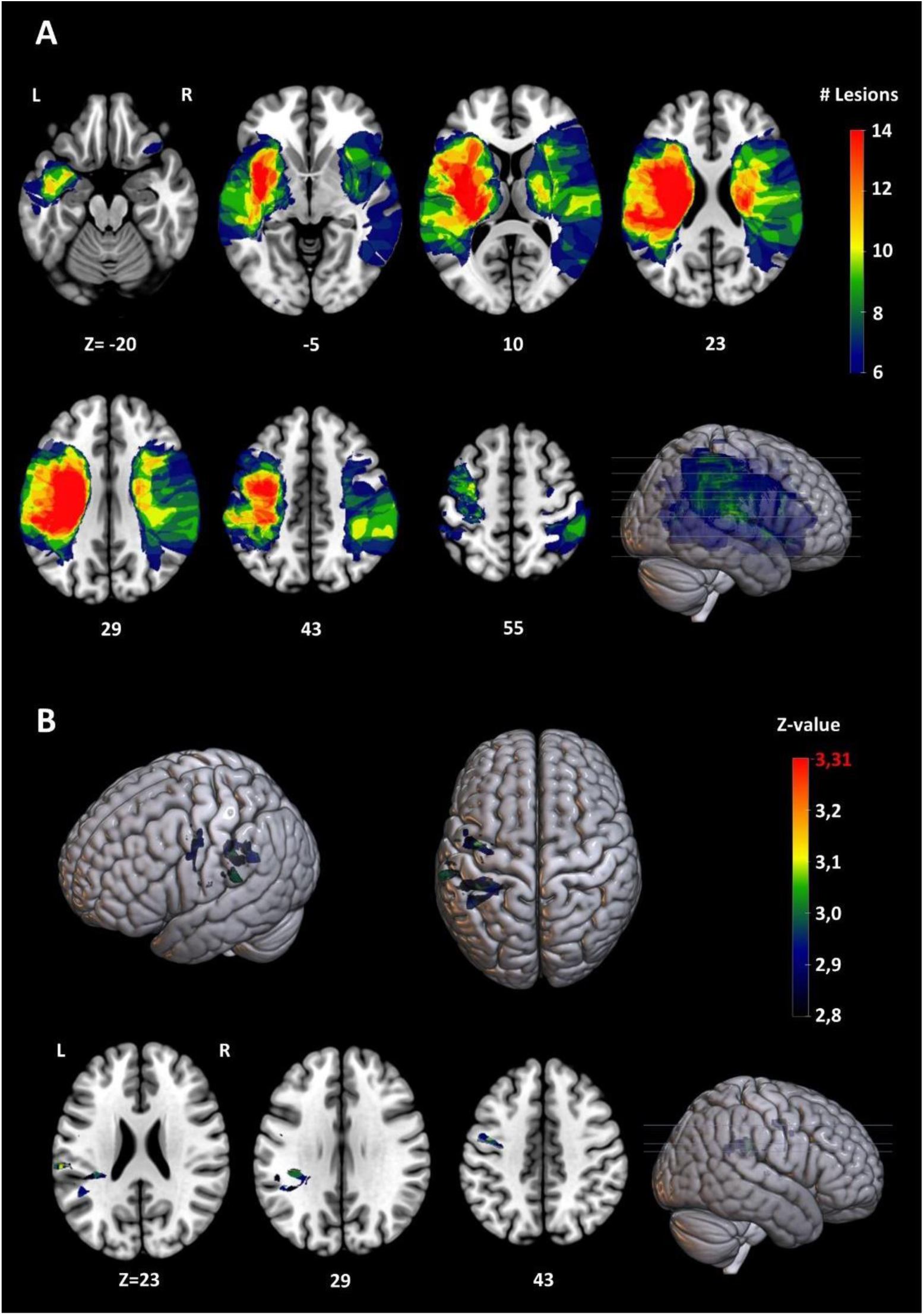
A: Brain lesion coverage map for voxels damaged in at least six patients; The color of each voxel represents the number of patients whose lesions covered the voxel. MNI z coordinates of the axial sections are reported. B: Lesion locations associated with the poor early recovery profile; Top row, 3D view of clusters presenting Z-values close to the FWER corrected significance threshold. *Bottom row,* axial sections crossing the same clusters. FWER corrected unilateral α = 0.05 threshold was Z=3.31; maximal significance value found was Z=3.29. MNI z coordinates of the axial sections are reported.

## 4. Discussion

### 4.1 The importance of early recovery in predicting the final outcome

In the present exploratory monocentric study conducted on a small cohort of stroke patients with moderate to severe impairment on admission we investigated whether the functional outcome upon discharge was predicted by the FIM gain during the first three weeks of SR (3wFIMgain). Patients’ outcomes were the functional level at discharge (DFIM) and the total amount of functional gain observed during the SR (DFIM gain).

We found that a value of 3wFIMgain>19 points successfully predicted (AUC=0.86, sensitivity=83.3%, specificity= 81.1%) which patient would present a clinically relevant functional gain in response to rehabilitation (DFIM gain>49). Using this single value to predict individual DFIM gain, we obtained a level of prediction comparable to that of more complex models based on multiple variables^8,9,32^ (R^2^=0.44). Because the DFIM gain is a direct estimate of rehabilitation effectiveness, the 3wFIMgain could identify patients who retained good recovery potential despite a moderate to severe initial impairment. Moreover, the finding that, unlike the admission FIM^3^ (AFIM) the 3wFIMgain predicts the DFIM gain, suggests that measures of early recovery might improve prognostic models because they account for the portion of outcome uncertainty that is due to the individual response to rehabilitation and is not fully predicted by the initial impairment.

Confirming these assumptions, we show that combining the 3wFIMgain to the AFIM in a linear model improves the efficacy predicting the DFIM (ΔR^2^=0.21). This two-predictor model also reaches the prediction capability of more complex multivariable models^3,8,9^ (R^2^=0.77). Importantly, if similar performances will be confirmed in alternative larger cohorts, our predictive model could be easily introduced in the daily clinical practice because it would be highly effective while remaining simple and based on readily available data^2,7^. And yet, it would still offers large margins of improvement, as additional factors might be identified and combined with the 3wFIMgain, or alternative nonlinear models could be tested.

Despite these promising features, our most crucial results concerned only a subgroup of 49 patients requiring a minimum length of stay in SR of 6 weeks (LOS, median = 81 days) to properly estimate the predictive power of the 3wFIMgain. Nevertheless, within the full cohort (N=81), the poor and good early recovery (PER/GER) groups showed no difference in the AFIM and NIHSS at stroke onset, suggesting that early recovery provided critical predictive information independent from the patient’s initial disability. This indicates that patients with GER are likely to achieve a favorable outcome independently from their condition on admission to SR, further highlighting the importance of combining measures of early recovery and initial disability to predict discharge outcomes.

While the advantage of integrating measures of early recovery in predictive models may seem obvious, it has remained relatively unexplored^9,10^. Tokunaga et al. (2017) found in a large cohort of stroke patients that adding the gain of the motor and cognitive FIM components obtained in the first month of SR to a four variables model improved considerably its performance in predicting the motor FIM gain at discharge. Koyama et al. (2005) developed a logarithmic model based on the FIM gain obtained in the first two to six weeks of SR and obtained highly reliable predictions of monthly FIM values until discharge in a small cohort of stroke patients. In the present study we confirmed the validity of the FIM gain obtained at even earlier stages of SR (three weeks) as a powerful prognostic predictor for patients with moderate to severe impairment and explored more in detail its relation to other common predictors.

One possible explanation for the limited use of early recovery measures in prognostic models might be that when using information acquired with a delay from admission, the prediction is postponed and therefore less useful. However, predictions based on the first three weeks of SR are still relevant because, while the average length of stays across hospital worldwide may vary, it frequently outlasts a three-week period for patients with severe-moderate deficits, whose prognosis is the most uncertain^16–19^. Thus, this period still represents a valuable time window to provide prognostic prediction that may be precious to better adapt the rehabilitative project to the patient’s profile of potential recovery.

### 4.2 The determinants of early recovery

We investigated whether PER or GER patients differed regarding clinical characteristics that might have influenced their early recovery. We expected the PER to be associated with older age^3,33^, a higher level of comorbidity^34,35^ and complications^36^, longer onset to rehabilitation admission delays^8,33^ (OAD), and worse levels of cognitive FIM on admission^3,9^ (AFIMc). A higher rate of aphasia^37^ and/or dominant-lateralized sensori-motor hemisyndrome^38^ was also anticipated.

Despite small trends in the expected direction for most indicators (AFIM, AFIMc, NIHSS, OAD, CIRS, complications, presence of aphasia/dominant hemisyndrome), we found no significant difference between the two groups. We also confirmed that NIHSS and AFIM are not major determinants of early recovery in patients with moderate to severe disability, indicating that the information contained in the 3wFIMgain is a necessary complement to those provided by measures of initial impairment. Therefore, when tested individually, these variables do not seem to impact early recovery. However, because post-stroke recovery depends on a combination of factors, some of these variables might reveal their effect if combined in a multivariate model.

Overall, our results support the 3wFIMgain as a single practical indicator to synthetize complex clinical information resulting from the interaction of multiple concurring factors.

### 4.3 Lesion locations associated with poor early recovery

As a supplementary investigation, we conducted a VLSM analysis on the full cohort to investigate lesion locations associated with the PER. Three clusters in the left hemisphere reached near-threshold significance: white matter deeply to the temporoparietal junction (TPJ), intersecting the superior longitudinal fasciculus (SLF), supramarginal gyrus (SMG) and precentral gyrus (PCG).

The SLF is a white matter bundle connecting a complex homolateral fronto-parieto-temporal associative network^39^. The left TPJ and SMG are associative areas connected with multiple homo– and contra-lateral brain regions^40^. All these structures are involved in high-order cognitive functions like language, attention, complex reasoning, etc.^39–41^. Thus, the association between their damage and PER suggests that impairments of complex cognitive functions reduce early recovery^3,42^. Specifically, a lesion to these associative hubs would simultaneously affect multiple cognitive domains possibly resulting in a synergic detrimental effect. Our results pointing to the PCG (but not the cortico-spinal tract) suggest that disrupted integration of complex sensorimotor input and motor learning^43^, rather than simple interruption of voluntary motor control transmission, has a worse impact on plasticity underlying early recovery.

Although our VLSM results did not reach statistical significance, we tested the influence of lesion-induced deficits, without accounting for other clinical factors like age, comorbidities, OAD, etc. Therefore, while among possible stroke-induced disabilities, dominant hemiparesis, aphasia and cognitive dysfunctions may represent those with the greatest impact on early recovery, their effect alone is not sufficient to determine a PER, but probably needs to interact with other concurrent factors.

### 4.4 Limitations

First, in line with the preliminary nature of this work, our predictive analyses were conducted on a subgroup of 49 patients with long rehabilitation stay. This cohort is relatively small considering the high clinical variability among stroke patients, which probably yielded low statistical power. Nonetheless, we revealed large effects replicating previous literature for the predictive power of AFIM.

Second, we conducted a monocentric study without validating our models. It would be crucial to externally validate our results in larger cohorts and multiple centers to confirm our findings and allow their generalization to the stroke population.

Third, although we analyzed patients with AFIM<100, ceiling effect was still present (Kwon et al. 2004). Consequently, the linear model for the prediction of DFIM tended to overestimate outcomes for patients with AFIM>80 approximately (figure 3c). However, the addition of the 3wFIMgain to the model reduced this overestimation (figure 3b vs 3c), showing at the same time a better performance within the AFIM range between 17 and 80 points, where outcomes are usually uncertain^5,6^. Nevertheless, this indicates that our models leave the margin for further improvement.

Finally, residuals of the two-predictor model of DFIM were more variable for the lower range of the predicted DFIM (≲80, figure 3c), meaning that predictions of severe disability at discharge were less reliable than those of moderate-mild disability. However, this aspect was partially corrected by the addition of 3wFIMgain to the model (figure 3b vs 3c).

### 4.5 Conclusions

Our study provides considerable base evidence that measures of functional early recovery might constitute a practical and effective way to synthetize complex prognostic information in a single and easily available variable that would allow for the development of more precise, yet simple prognostic models to help clinicians in predicting functional outcome and plan SR of individual patients.

Our VLSM findings suggest that the deep white matter of the temporo-parietal junction, supramarginal and precentral gyrus in the left hemisphere are among the locations whose damage is more likely associated with a poor response to the first three weeks of SR. However, their effect alone is not sufficient to determine such a poor response but might require the interaction with other specific clinical factors. Our findings highlight the need to further investigate the predictive validity of measures of early recovery through external validation of our results in larger multicentric cohorts.

## Supporting information

supplement analysis

## Data Availability

All data produced in the present study are available upon reasonable request to the authors

## Acknowledgments

This work was supported by a grant from the Swiss National Science Foundation to LS (#32513B_212616) and AT (#320030_188737), as well as by the QuadriMed fund, and the Fondation Pierre Mercier pour la science. We thank Franziska Peier and Naima Mory for their help in data collection and Fiona Chesnel for her help in lesion processing.

