## supplement analysis for "Early recovery during stroke rehabilitation predicts the functional outcome at discharge: a retrospective clinical study"

***Supplementary results***

***Association of the early recovery profiles with outcomes***

Within the subgroup of patients with a LOS ≥ 6 weeks (median= 81 days), patients with PER (N=32) or GER (N=17) showed significant differences in terms of DFIM (Mann-Whitney U test, $p<0.01$, mean difference=28) and DFIM gain (Welch's t(38.0), $p<0.01$, Cohen's d=1.72) with the two outcomes being better in the GER group. The two groups had similar AFIM (Mann-Whitney U test, p=0.34, mean difference=-6.1).

The Euclidean distance from the PER group showed a positive low/moderate correlation with the DFIM (Spearman's rho(49) =0.39, $p<0.01$) and a moderate positive correlation with the DFIM gain (Pearson's r(49) = 0.60, $p<0.01$). Within the same subgroup of patients with a LOS ≥ 6 weeks, the AFIM showed a strong positive correlation with the DFIM (Spearman's rho(49) =0.81, $p<0.01$), but no significant correlation with the DFIM gain (Spearman's rho(49) =0.09, p=0.52). Similarly, the NIHSS showed a moderate negative correlation with the DFIM (Spearman's rho(35) =-0.47, $p<0.01$), but no significant correlation with the DFIM gain (Spearman's rho(35) =-0.01, p=0.95). The NIHSS was available in only 35 patients of the subgroup. The lack of correlation of AFIM and NIHSS with the DFIM gain was partially influenced by a ceiling effect due to the portion of patients with high functional level on admission (AFIM >80).
